# Sexuality and respiratory outcomes in the UK: disparities, development and mediators in multiple longitudinal studies

**DOI:** 10.1101/2025.02.07.25321888

**Authors:** Evangeline Tabor, Dylan Kneale, Praveetha Patalay

## Abstract

**Background:** Limited international research suggests sexual minority individuals, in particular sexual minority women, may experience worse asthma outcomes than their heterosexual peers. However, little research has explored if a similar disparity is present in the UK.

**Methods:** Pooling data from five UK longitudinal studies (total N=86,024, 4,860 (5.65% sexual minority) we examined asthma and lung function by sexuality. To explore the impact of ageing on disparities we compared asthma by sexuality at two time points approximately 10 years apart. To help understand potential causal mechanisms we used respondents aged under 18 as a negative control. Finally, we conducted a mediation analysis to examine the extent to which smoking accounts for asthma disparities.

**Findings:** Sexual minority respondents were more likely to report asthma (RR 1.43 [95% CI 1.26 to 1.62]) and poor lung function (RR 1.50 [95% CI 1.20 to 1.87]). Sexual minority women (RR 1.51 [95% CI 1.30 to 1.76]) and bisexual respondents (RR 1.75 [95% CI 1.35 to 2.26]) were more likely to report asthma than their heterosexual counterparts. Rates of asthma did not vary by sexuality in <18s and disparities increased between time points in adults supporting the hypothesis that disparities emerge after childhood and widen over the lifecourse. Smoking only partially mediated (proportion mediated 1%) the relationship between sexual minority status and asthma.

**Conclusions:** This analysis adds substantially to our understanding of how and when asthma and lung function disparities by sexuality in the UK emerge, as well as evidencing the limited role of smoking.

**Key Messages:**

- Our research question was whether respiratory health, in particular asthma, varied by sexuality and how it patterned across the lifecourse.
- We found that sexual minority people were at greater risk of asthma and poor lung function than heterosexual people and this disparity emerges and widens in adulthood.
- Respiratory disease can impact quality of life and mortality risk and understanding patterns and mechanisms of disease in the LGBTQ+ populations can allow us to address health inequalities.

## Introduction

Asthma is a common chronic respiratory condition, estimated to affect 262 million people worldwide, which causes coughing and breathlessness and, in severe cases, increases mortality risk (1–3). Evidence from the USA indicates there are inequalities in asthma by sexuality, with some multi-outcome studies in healthcare data suggesting similar patterns in England (4–6). This inequality has been attributed to a number of mechanisms which crystalise in adulthood including differential rates of smoking, physical activity and obesity, migration to urban centres, psychosocial stress resulting from structural and interpersonal discrimination, and healthcare access/experience (4,7–9).

However, to date, there has been no study focussed on asthma inequalities by sexuality in the UK; it’s longitudinal development, underlying functional measures, age at emergence and potential mechanisms. The existing England-only studies are primarily concerned with multi-disease burden and do not contextualise specific disease outcomes nor test potential mechanisms (5,6). As asthma and many related mechanisms are chronic and cumulative, the patterning of inequality may change or even widen as individuals age and relevant biopsychosocial exposures accumulate. As a result, there is a clear need for an expanded evidence base for asthma in sexual minority people in population-based studies, in particular focusing on outcomes over the life course and potential mediators.

This current study aimed to expand the evidence for the relationship between sexuality and asthma in the UK while adding substantially to our understanding of underlying mechanisms. We examine the patterning of the disease, particularly by sex, sexual identity, and age, and examine outcomes longitudinally. This analysis makes use of a negative control outcome to evidence the timing of the emergence of asthma disparities and provide more robust inference about causality. We also explore outcomes in an objective measure of lung function, which also helps unpack the potential ascertainment bias introduced by differential access to healthcare services. Finally, this analysis also seeks to examine and quantify the role of smoking – which has been previously identified as a potential factor - as a mediator of the relationship between sexuality and asthma (4).

## Methods

### Design

We use a co-ordinated individual level data analysis strategy across multiple longitudinal population-representative studies in the UK, this multi-dataset approach helps with sample sizes and power, which would be limited for the sexual minority group if using only one data source. Given the proposed mechanisms begin in adolescence and crystallise in adulthood, there should be no differences by sexuality observed in childhood- we test this by conducting a negative control outcome analysis of asthma outcomes in childhood. To examine whether inequalities widen with age we examine differences by sexuality in the same people approximately ten years apart. Lastly, mediation analysis with smoking as a mediator of observed inequalities is tested. A preregistered protocol for this study is available at the Open Science Framework (https://osf.io/mr6yb/).

### Data and participants

The study utilised data from five longitudinal population studies: the Millennium Cohort Study (MCS), Next Steps, the British Cohort Study (BCS70), Understanding Society: the UK Household Longitudinal Study (UKHLS), and the English Longitudinal Study of Ageing (ELSA)(10–14). See the supplementary materials for more information about each dataset.

Data sources were included if they: (a) measured sexual identity; (b) collected data longitudinally from the same individuals (i.e. birth cohort or panel studies); (c) collected information on asthma and/or lung function; and d) were sampled in the UK using probabilistic and representative methods.

### Variables

#### Sexuality

All respondents were asked for the sexual identity label which best described them at age 17 in MCS (2018), wave 8 in ELSA (2016-17), at multiple waves in UKHLS, and at age 42 (2012) in BCS70. Further detail on the coding of this variable is available in the supplementary materials (15). In all analyses, “Heterosexual” is the reference category.

#### Asthma diagnosis

For each study, a binary variable of ever diagnosed with asthma is used. Study member reported diagnosis is used for ELSA, UKHLS, BCS70, and Next Steps and study member and parent report is used for childhood sweeps in MCS. Where available asthma diagnosis at most recent full wave is used, however for MCS and Next Steps the COVID-19 sub-sample wave collected in 2020-2021 is used.

#### Lung Function

A measure of lung function (spirometry) was collected in 2010-12 and 2011-13 in UKHLS and 2012- 13 in ELSA. Lung function was measured using the forced expiratory volume in the first second of exhalation (FEV1). FEV1% or the percentage of the expected FEV was considered as a continuous outcome and a binary outcome was created where outcomes below 80% will be considered obstructed (16,17). Most recent response for each individual is used where more than one measure was available.

#### Smoking status

Smoking status was operationalised as a two-category variable comprising “Never smoker” and “Ever Smoker” assessed at the same sweep as the asthma diagnosis.

#### Covariates

In addition, the following were included as covariates: year of birth, a binary sex variable comprising “Male” and “Female”, binary ethnicity variable comprising “White” and “Other”, a binary education variable comprising “Degree or equivalent” and “All else”, a four category employment variable comprising “Managerial /Admin /Professional”, “Intermediate”, “Manual/routine”, and “Other/Never worked”, and variable indicating study.

### Statistical Methods

First, we pooled data from all studies to examine asthma diagnosis by sexuality. This analysis used the responses at the most recent full sweep of each study where available. An Individual Participant Data (IPD) meta-analysis was conducted using the pooled data (18). Binomial logistic regression was conducted to examine the impact of sexuality on the binary outcome of reported asthma diagnosis.

Secondly, to examine whether differences in asthma vary due to aging, we conducted binomial logistic regression of asthma diagnosis by sexual minority status for each study at two sweeps: the most recent sweep of the survey, and the sweep collected approximately ten years prior. This analysis used data from MCS at age 7 (2008) and MCS COVID-19 sweeps (2020/21), UKHLS at wave 1 (2009-11) and wave 11 (2019-21), BCS70 at age 34 (2004) and age 46 (2016), × ELSA at wave 4 (2008-9) and 9 (2018-19). In this analysis, childhood asthma was considered a negative control outcome as no difference in asthma outcomes was predicted at this age by adult sexual identity.

Thirdly, we used pooled data to examine lung function outcomes. Linear and logistic regression was conducted in a sub-sample comprising UKHLS wave 2 (2010-12) and 3 (2011-13) and ELSA wave 6 (2012-13) to explore the relationship between sexual minority status and FEV1% outcomes. Due to differences in instruments used, the studies were analysed separately and then outcomes recategorized into a binary for a pooled analysis.

The final analysis used pooled data from all studies to examine whether differences in reporting asthma between sexual minority respondents and their heterosexual peers are mediated by smoking status. This analysis used the responses at the most recent full sweep of each study where available. A mediation analysis was performed using the gformula package in Stata (using 250 bootstrap replications (19)) using logistic regression models to estimate the direct, indirect, and total effect. The mediation model was adjusted for confounding by sex, age, ethnicity, employment and education. This analysis used pooled data of all respondents aged 18 and over.

The pooled analysis approach was selected due to small sample sizes in the individual studies and to overcome issues of power in previous research in this area. In addition, the greater N allows for the examination of differences by specific sexual identities. All analyses were conducted with no adjustment and with confounder adjustment for sex, age, ethnicity, study member (/parent) income and education level, and study.

Stratified analysis was conducted by sex in all datasets, and additionally by gender identity in MCS. In addition, where sample size allowed, pooled analysis by individual sexualities were conducted.

Each of the datasets have a slightly different design so where possible we used weights to account for survey design, non-response and attrition in all of the datasets.

### Sensitivity analyses

To answer the first two research questions and their sub-questions, a secondary two-stage IPD analysis was also conducted. In the two stage meta-analysis results from each study were pooled using a random effect (IPD) meta-analysis and formally tested for heterogeneity across studies (I^2^ statistic).

A further sensitivity analysis was run where study members who report a “Mainly Heterosexual” identity were recategorized as “Sexual minority” in MCS.

## Results

We conducted analyses using 5 longitudinal studies, with 86,024 participants in total (including 46,989 [54.62%] women and 39,035 [45.38%] men). Of the participants, 4,860 (5.65%) were sexual minority. Participants ranged from age 16 years to 90+ years. 67,844 (78.87%) of participants were White, 17,504 (20.35%) non-White, and 676 (0.79%) were missing ethnicity. Individual study samples ranged from 4,390 in Next Steps to 53,811 in UKHLS. Descriptive statistics for exposures, outcome, and covariates are presented in *Table 1*.

**Table 1:**
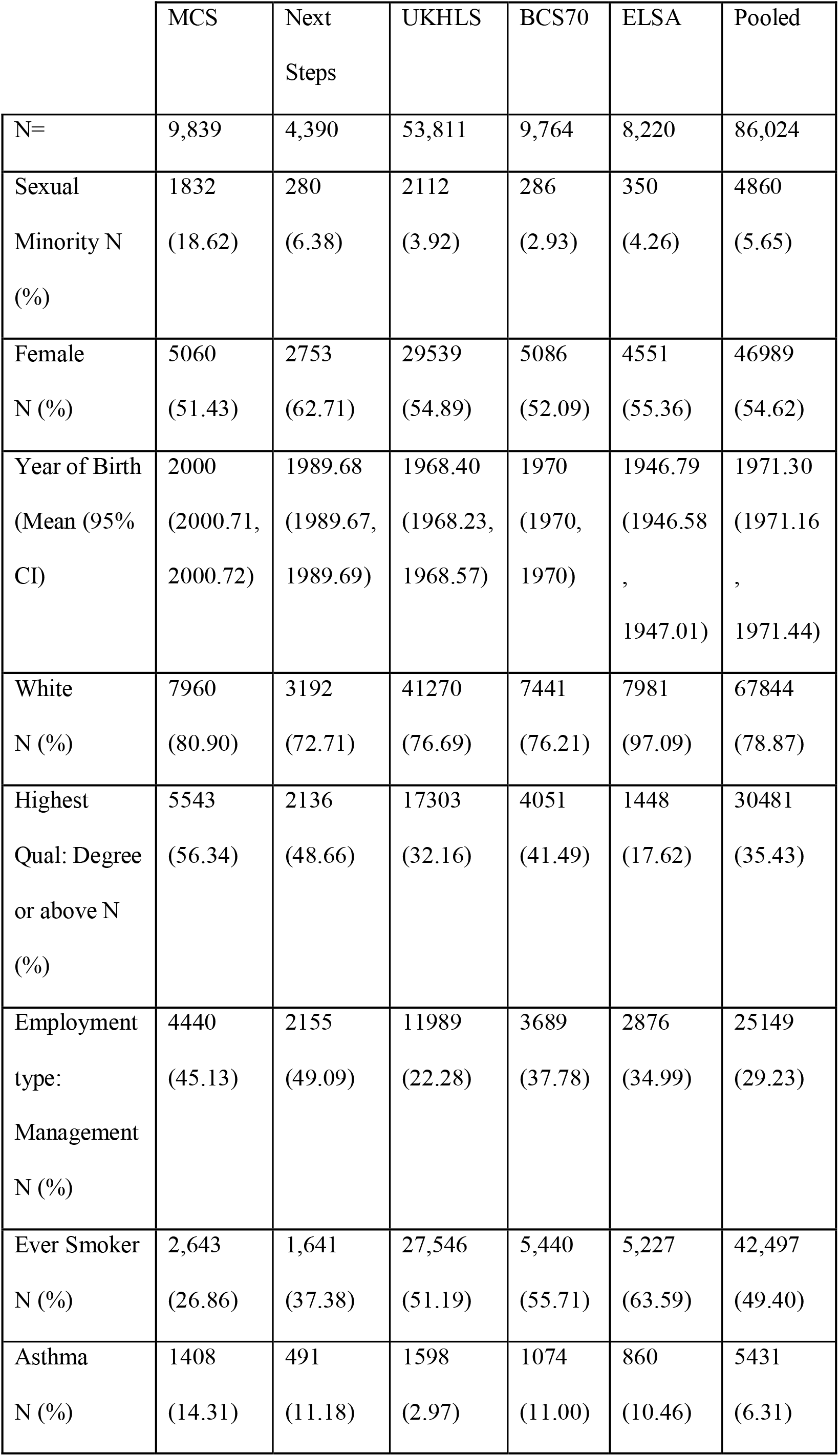
Description of asthma analysis sample.

### Asthma outcomes by sexual minority status

10.14% of sexual minority respondents reported receiving a diagnosis of asthma compared to 6.12% of heterosexual respondents. Estimates adjusted for confounders indicate that sexual minority adults were more likely to report asthma than heterosexual adults (RR 1.43 [95% CI 1.26 to 1.62]) (see Figure 1).

**Figure 1:**
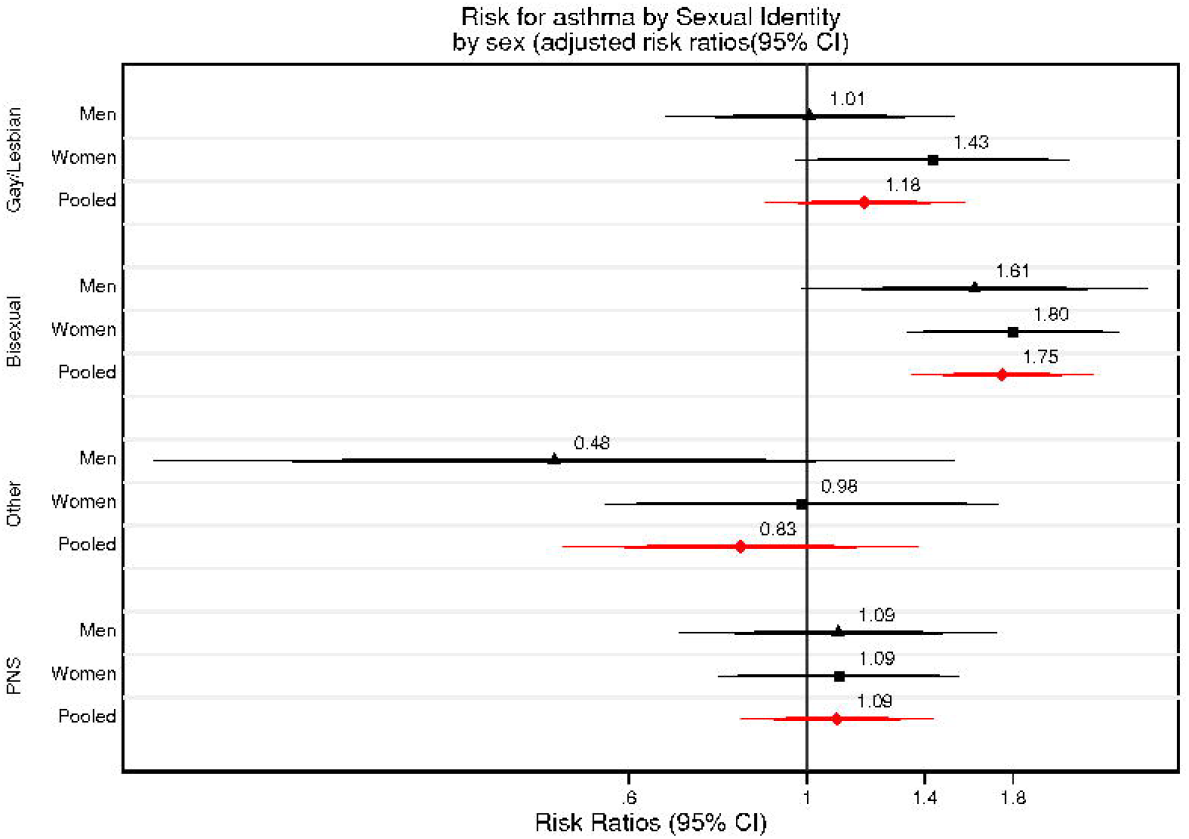
Association between Sexual Minority status and Asthma report a) by study and b) by sex, adjusted risk ratios

11.64% of sexual minority women reported asthma compared to 6.38% of heterosexual women, while 7.88% of sexual minority men and 5.83% of heterosexual men also reported asthma. Pooled adjusted estimates indicated that sexual minority women had a higher likelihood of asthma than heterosexual women (RR 1.51 [95% CI 1.30 to 1.76]). Sexual minority men also had a higher likelihood of asthma than heterosexual men, however the association was weaker (RR 1.29 [95% CI 1.02 to 1.62]) (see *Figure 1*).

When examining sexual identity subgroups, adjusted estimates indicated that gay/lesbian adults (RR 1.18 [95% CI 0.88 to 1.57]) and bisexual adults (RR 1.75 [95% CI 1.35 to 2.26]) had a higher likelihood of asthma than heterosexual adults (see *Figure 2*). However, Other and Prefer Not To Say adults had equal or less likelihood of reporting asthma than heterosexual adults.

**Figure 2:**
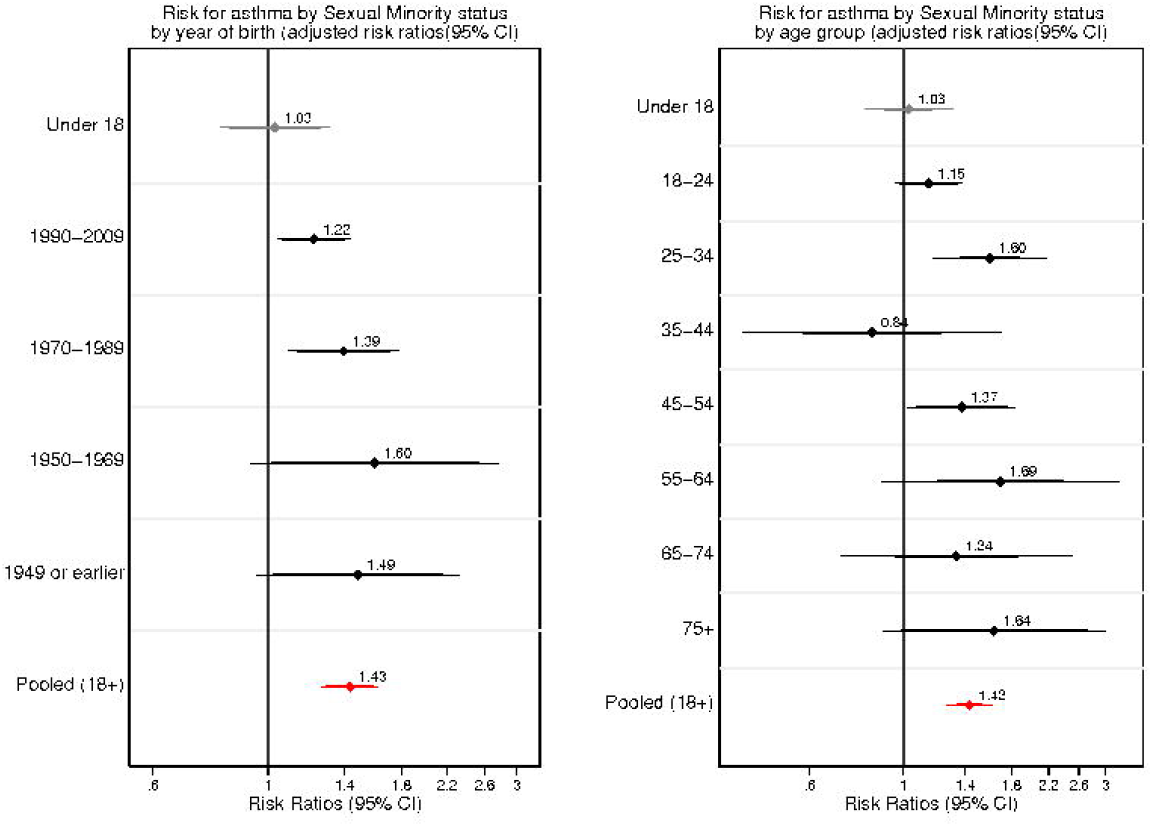
Association between Sexual Identity and Asthma report by sex, adjusted risk ratios.

Similar results are observed when we conducted a sensitivity analysis using a 2-stage meta-analysis (see Supplementary *Figures 1 and 2*).

### Asthma over time

The likelihood of asthma increased over time in adulthood (T1: RR 1.06 [95% CI 0.91 to 1.22], T2: RR 1.30 [95% CI 1.10 to 1.53]). No difference was observed in the childhood cohort: MCS.

When stratified by sex, the likelihood of asthma in sexual minority women compared to heterosexual women increased over time (T1: RR 1.12 [95% CI 0.93 to 1.33], T2: RR 1.37 [95% CI 1.12 to 1.68]), whereas in sexual minority men it decreased over time (T1: RR 1.40 [95% CI 1.11 to 1.76], T2: RR 1.21 [95% CI 0.90 to 1.65]).

### Negative control analysis

Across all age groups over 18 years, sexual minority respondents had a higher likelihood of asthma than their same-age heterosexual peers (see *Figure 3*). Risk compared to heterosexual adults also appeared to increase by age or year of birth within different adult age bands.

**Figure 3:**
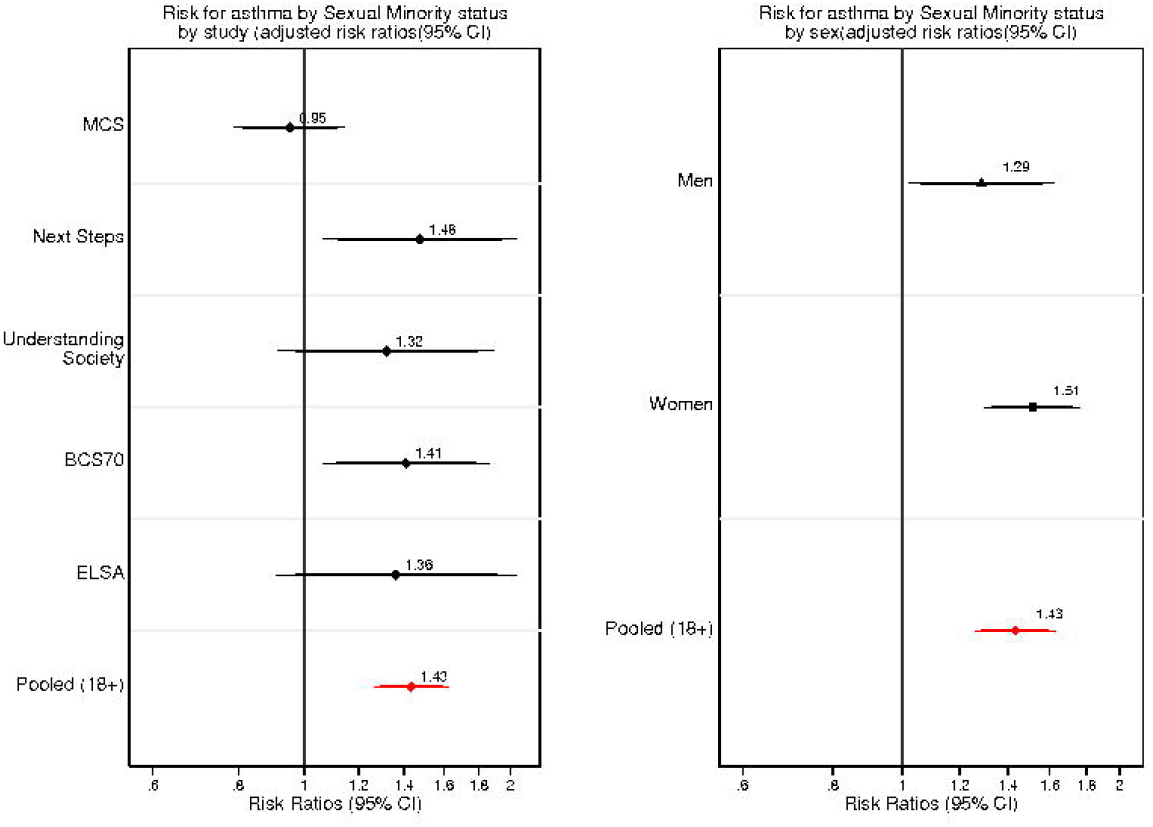
Association between Sexual Minority status and Asthma report a) year of birth and b) by age, adjusted risk ratios

In the negative control analysis, we stratified by age and found that among respondents under 18 there was no difference in risk by sexuality (see Figure 4). In addition, there was no difference at time points ten years apart and no significant change over time in the under 18 sample (see supplementary *Figure 3*).

**Figure 4:**
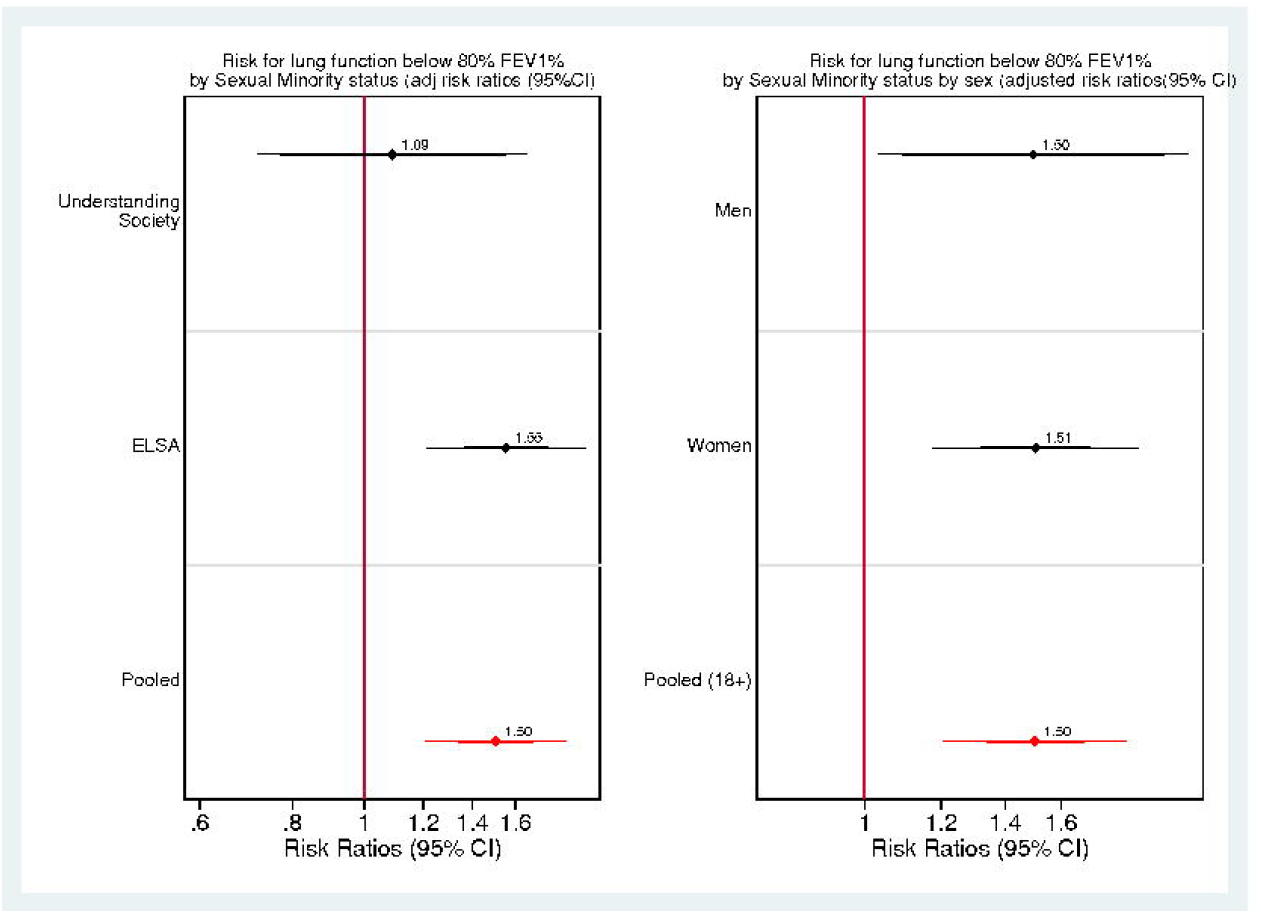
Association between Sexual Minority status and lung function below 80% FEV1% by a) study and b) sex, adjusted risk ratios.

In a sensitivity analysis, regardless of categorisation of “Mainly Heterosexual” respondents as “Heterosexual” or “Sexual Minority” results remained the same (see supplementary *Table 6*)

### Lung function

Pooled adjusted estimates indicated that sexual minority adults were more likely to report poor lung function than heterosexual adults (RR 1.50 [95% CI 1.20 to 1.87] (see *Figure 4*).

When examined using continuous FEV1%, sexual minority adults recorded poorer lung function than heterosexual adults (see appendix Figure 8-4). However, this varied substantially by study (e.g. UKHLS: -0.33 [95% CI -1.27 to 0.62], ELSA: -4.86 [95% CI -8.40 to -1.32]), likely due to differences in study sample age. No pooled result was calculated as the continuous lung function measures were measured using different machines.

### Smoking as a mediator

Around half of sexual minority adults (49.7%) and heterosexual adults (51.84%) reported ever smoking. Adjusted risk ratios indicate that sexual minority adults had higher likelihood of smoking than heterosexual adults (RR 1.07 [95% CI 1.03 to 1.12]).

In the pooled analysis, the natural direct effect between sexual minority status and asthma was estimated at 0.03 [95% CI 0.02 to 0.04]. The natural indirect effect between sexual minority status and asthma through smoking status was estimated at 0.0003 [95% CI -0.002 to 0.003]. The total causal effect was estimated at 0.03 [95% CI 0.02 to 0.04]. From these results, smoking status mediates only a very small part of the association between sexual minority status and asthma (Proportion Mediated: 1%).

## Discussion

Overall, our analysis found UK sexual minority adults are at a higher risk of asthma than heterosexual adults. Sexual minority women and bisexuals are at particular risk compared to their heterosexual counterparts. Sexual minority people under the age of 18 had a similar likelihood of asthma than their heterosexual peers, indicating that differences in asthma may emerge in adulthood due to specific mechanisms that become more relevant after childhood (e.g. discrimination on the basis of sexuality, alcohol and tobacco use). We also found that difference in risk of asthma widened over time supporting the hypothesis that the difference is at least in part driven by the accumulation of exposure to social and-environmental factors (20). Sexual minority adults were also more likely to report low lung function. Finally, we found that the difference in asthma risk between sexual minority and heterosexual adults is only partially mediated by smoking indicating further investigation of mediating factors is needed.

### Asthma in sexual minority adults

This analysis found sexual minority adults are at a higher risk of asthma than heterosexual adults. We found sexual minority women have a particularly high risk for asthma compared to heterosexual women. These findings are particularly notable given asthma is more common in women in the general population so may indicate that sexual minority women are at a particularly high risk (2). While sexual minority women in this analysis were more likely to smoke, as the analysis shows, this does not fully account for the relationship.

Additionally, bisexual adults had a higher risk for asthma than heterosexual adults. These findings are consistent with literature which has identified bisexuals as a group at particular risk for a number of mental and physical conditions. As well as the broader processes effecting sexual minority people, bisexuals are subject to experiences of biphobia and exclusion from both heterosexual and sexual minority communities which may result in increased distress, poorer health behaviours, and therefore worse health outcomes (21–23).

Sexual minority people are less likely to seek medical treatment so relying on a diagnosis of asthma may result in an undercount (24). To account for possible ascertainment bias, as well as to compare the severity and any underlying functional outcomes, we also examined lung function. We found that sexual minority adults had worse lung function outcomes than heterosexual adults. While lung function is indicative of more than just asthma, it indicates that not only are sexual minority people more likely to experience symptoms resulting in a diagnosis of asthma, but that lung functionality is also significantly decreased. This points to underlying issues that may not be captured by looking only at downstream outcomes.

### Mechanisms

In previous research, asthma disparities in sexual minority people has been attributed to higher prevalence of risky health behaviours, higher urbanity, and increased psychological stress due to experiences of interpersonal and structural discrimination (9). The majority of these biological, psychological, environmental and social processes are cumulatively embodied over time and interact across the lifecourse resulting in patterns of exposure and susceptibility (20,25). We found sexual minority people under the age of 18 had similar likelihood of asthma than their heterosexual peers. However, in adults, there was a difference in risk which widened over a ten-year period, regardless of respondent age. These results support the hypothesis that differences in asthma emerge in adulthood and are in part driven by the accumulation of exposure to socio-environmental factors. Differences only began to emerge in young adults aged 18 to 34 which may indicate a potential window for intervention before disparities begin to crystalise.

Finally, differences in smoking are highlighted throughout the literature as a primary driver of sexual minority disparities in asthma (4). However, while we found that sexual minority adults were more likely to have ever smoked, the difference in asthma risk was only partially mediated by smoking. This research highlights the importance of future investigation of other potential causes, for example, urbanity, relationship with obesity, access to medical services, and the consequences of chronic stress.

### Strengths and Limitations

This analysis presents results from five large nationally representative longitudinal population studies. The study adds key evidence on risk for asthma and underlying lung function in the UK population, and incorporates novel outcomes such as a measure of lung function and exploration of change over time.

However, this paper has a number of important limitations. Due to non-random attrition, the sample is likely healthier and wealthier than the general population. This is a particular issue in the older studies where attrition due to illness or mortality may result in an overall survivor bias. Sexual minority populations are subject to histories of structural discrimination and neglect and have well documented risk of poor mental health and HIV/AIDS which may result in disproportional levels of attrition due to associated disability or mortality.

A further limitation of this analysis was the lack of measures of e-cigarette use which plays a role in contemporary patterns of smoking (26).

## Conclusion

Asthma can impact lifelong quality of life, mortality risk, and add to multi-morbidity and polypharmacy burdens. Asthma in sexual minority people is chronically understudied in the UK context despite the potential impact on LGBTQ+ people’s health across the life course. We found sexual minority people, in particular sexual minority women and bisexuals, were at higher risk of asthma than heterosexual people. This difference in asthma appears to emerge in adulthood and widen across time. Finally, while much attention has been given to higher prevalence of smoking among sexual minority people, we found smoking only partially explained the asthma disparity so call for further research into other potential causes.

## Ethics approval

Ethics approval for this analysis was given by the Institute of Education Ethics Committee, University College London (approval number: N/A). All data was collected with approval from the corresponding ethics committees (further detail in Supplementary).

## Funding

This work was supported by the Economic and Social Research Council - Biotechnology and Biological Sciences Research Council Soc-B Centre for Doctoral Training (ES/P000347/1) to ET.

## Data Availability

All data is publicly available from the UK Data Service (https://ukdataservice.ac.uk/).

## Supplementary data

Supplementary data are available at IJE online

## Author contributions

ET conceptualised and designed the study, designed the analytical strategy, conducted the analysis, and prepared the manuscript. DK and PP reviewed and edited the manuscript and supervised ET.

## Conflict of interest

None declared.

## Notes

### Competing Interest Statement

The authors have declared no competing interest.

### Clinical Protocols

https://osf.io/mr6yb/

### Author Declarations

All data is publicly available from the UK Data Service (https://ukdataservice.ac.uk/).

