## Supplementary Materials for "Sexuality and respiratory outcomes in the UK: disparities, development and mediators in multiple longitudinal studies"

### Research in context systematic search terms:

PubMed terms example:

(("asthma"[Title/Abstract] OR "respiratory"[Title/Abstract] OR "wheezing"[Title/Abstract] OR "chronic obstructive pulmonary disease"[Title/Abstract] OR "COPD"[Title/Abstract] OR "lung"[Title/Abstract]) AND 1000/01/01:2024/12/12[Date - Publication] AND (("sexual minority"[Title/Abstract] OR "lgbt"[Title/Abstract] OR "sexuality"[Title/Abstract] OR "lesbian"[Title/Abstract] OR "gay"[Title/Abstract] OR "bisexual"[Title/Abstract])

### Studies:

All datasets are available through the UK Data Service.

#### Millennium Cohort Study (MCS)

The Millennium Cohort Study (MCS) is a large longitudinal birth cohort study of children born between September 2000 and January 2002 (1). 18,552 families were originally recruited across the UK, with 10,625 families responding at the most recent wave (2). There have been seven data collection sweeps at ages 9 months, 3, 5, 7, 11, 14, and 17 years to date (1). Children living in disadvantaged areas, and children from ethnic minority backgrounds were deliberately oversampled to allow analysis of these populations (1). Data collection consists largely of face-to-face interviews and paper questionnaires, but MCS has also collected self-completed questionnaires, activity monitor data, and objective health measures (1). Further detail is available elsewhere (1).

##### Supplementary Table 1: MCS data collection and sample achieved.^[1]^

| Sweep Year | Age at sweep | Interview type | Family sample achieved | Sample replenished? |
| --- | --- | --- | --- | --- |
|  | 9 months | Parent interview | 18,552 | - |
|  | 3 years | Parent interview, sibling interview, child assessment | 15,590 | 692 families – mostly returnees or new arrivals to UK. |
|  | 5 years | Parent interview, sibling interview, child assessment & measurement, teacher questionnaire | 15,246 | - |
|  | 7 years | Parent interview, cohort member assessment and measurement, cohort member self-completion, teacher questionnaire | 13,857 | - |
|  | 11 years | Parent interview, cohort member assessment and measurement, cohort member self-completion, teacher questionnaire | 13,287 | - |
|  | 14 years | Parent interview, cohort member assessment and measurement, member self-completion | 11,726 | - |
|  | 17 years | Parent interview, cohort member interview, child assessment and measurement, cohort member self-completion | 10,625 | - |

Ethics approval for the MCS study was obtained from the National Research Ethics Service Committee London— Central (reference 13/LO/1786). All parents gave consent for their children to participate and young people also provided verbal consent.

#### Next Steps

*Next Steps* (formerly the Longitudinal Study of Young People in England (LSYPE)) is a longitudinal study of around 16,000 people born in England in 1989/1990 (3). Participants were recruited in 2004 via schools in England when they were aged 13-14 years old (3). There have been eight full data collections sweeps to date at ages 14, 15, 16, 17, 18, 19, 20 and 25, with COVID-19 surveys at age 31 (3,4). Further detail is available elsewhere (3).

##### Supplementary Table 2: Next Steps data collection and sample achieved

| Sweep Year | Age at sweep | Interview type | Sample achieved | Sample replenished? |
| --- | --- | --- | --- | --- |
| 2004 | 14 | Parent and cohort member interview face to face | 15,770 | - |
| 2005 | 15 | Parent and cohort member interview face to face | 13,539 | - |
| 2006 | 16 | Parent and cohort member interview face to face | 12,439 | - |
| 2007 | 17 | Parent and cohort member interview face to face | 11,449 | - |
| 2008 | 18 | Cohort member interview via online, telephone or face to face. | 10,430 | - |
| 2009 | 19 | Cohort member interview via online, telephone or face to face. | 9,799 | - |
| 2010 | 20 | Cohort member interview via online, telephone or face to face. | 8,682 | - |
| 2015 | 25 | Cohort member interview via online, telephone or face to face. | 7,707 | - |

Next Steps was granted ethical approval for each sweep from 2000 by the National Health Service (NHS) Research Ethics Committee and all participants have given informed consent.

#### 1970 British Cohort Study (BCS70)

The 1970 British Cohort Study is a longitudinal birth cohort study of children born in a single week in 1970 in England, Scotland and Wales (5). Individuals born during this week who later immigrated to the UK were added to the sample up until the cohort reached adulthood (5). Data collection sweeps were conducted at birth, and at ages 5, 10, 16, 26, 30, 34, 38, 42, 46 with a further sweep planned at age 50 (5,6). While the majority of the data collection has been collected through face-to-face interviews, the study has also employed postal surveys and conducted a nurse visit at age 46 (6). Further detail is available elsewhere (5).

##### Supplementary Table 3: BCS70 data collection and sample achieved^[2]^

| Sweep Year | Age at sweep | Interview type | Sample achieved | Sample replenished? |
| --- | --- | --- | --- | --- |
|  | Birth | Mother interview, midwife questionnaire, medical examination | 17,196 | - |
|  | 5 years | Parent interview, health visitor questionnaire, cohort member assessments, medical examination and measurements | 13,135 | New arrivals in the UK included |
|  | 10 years | Parent interview, health visitor questionnaire, cohort member self-completion, cohort member assessments, medical assessment and measurements | 14,675 | New arrivals in the UK included |
|  | 16 years | Parent interview, medical assessment and measurements, cohort member self-completion, cohort member assessments, teacher questionnaires | 11,622 | New arrivals in the UK included |
|  | 26 years | Paper cohort member questionnaire | 9,003 | - |
|  | 30 years | Cohort member interview. | 11,261 | - |
|  | 34 years | Cohort member interview, cohort member assessment, parent and child module. | 9,665 | - |
|  | 38 years | Cohort member telephone interview. | 8.874 | - |
|  | 42 years | Cohort member interview, cohort member self-completion, cohort member assessment | 9,841 | - |
|  | 46 years | Cohort member interview, cohort member assessments, cohort member self-completion, nurse visit | 8,581 | - |
|  | 50 years | Cohort member interview, cohort member self-completion. | - | - |

BCS70 was granted ethical approval for each sweep from 2000 by the National Health Service (NHS) Research Ethics Committee and all participants have given informed consent.

#### Understanding Society: the UK Household Longitudinal Study (UKHLS)

Understanding Society: the UK Household Longitudinal Study (UKHLS) is a large longitudinal panel survey of approximately 40,000 households across England, Scotland, Wales and Northern Ireland (7). The survey consists of members of the British Health Panel Survey, a large General Population Sample recruited using a stratified, equal-probability sample of residential addresses, and ethnic minority boost samples (UKHLS, 2019). Household members are visited at a similar time of year each wave and data collection takes place over a 24 month period (7). The main interview is largely conducted face-to-face, although from the 2011-13 wave a proportion of respondents were interviewed by phone and from the 2015-17 wave a proportion undertake a web interview (7). The self-completion questionnaire was administered by paper, and then by computer starting from Wave 3 (7). Finally, a nurse visit was conducted in 2010-2012 with a subset of the General Population Sample and in 2011-13 with the British Health Panel Survey sample (8).

##### Supplementary Table 4: UKHLS data collection and calculated sample achieved ^[3]^

| Sweep Number / Year | Interview type | Adult sample achieved | Sample replenished? |
| --- | --- | --- | --- |
| 1 (2009-11) | Household questionnaire, individual interview, individual self-completion | 47,733 | Includes Ethnic Minority Boost Sample |
| 2 (2010-12) | Household questionnaire, individual interview, individual self-completion, nurse visit (subsample) | 50,665 | British Household Panel Survey Sample added (approx. 8,000 households) |
| 3 (2011-12) | Household questionnaire, individual interview, individual self-completion, nurse visit (BHPS subsample) | 45,601 | - |
| 4 (2012-14 | Household questionnaire, individual interview, individual self-completion | 43,326 | - |
| 5 (2013-15 | Household questionnaire, individual interview, individual self-completion | 40,920 | - |
| 6 (2014-16) | Household questionnaire, individual interview, individual self-completion | 33,733 | Immigrant and Ethnic Minority Boost Sample added (approx. 2,900 households) |
| 7 (2015-17) | Household questionnaire, individual interview, individual self-completion | 34,954 | - |
| 8 (2016-18) | Household questionnaire, individual interview, individual self-completion | 33,457 | - |
| 9 (2017-18) | Household questionnaire, individual interview, individual self-completion | 35,063 | - |
| 10 (2018-20) | Household questionnaire, individual interview, individual self-completion | 34,318 | - |
| 11 (2019-21) | Household questionnaire, individual interview, individual self-completion | 32,006 | - |

The University of Essex Ethics Committee has approved all data collection on Understanding Society main study and innovation panel waves, including asking consent for all data linkages except to health records. Approval for the collection of biosocial data by trained nurses in Waves 2 and 3 of the main survey was obtained from the National Research Ethics Service (Understanding Society - UK Household Longitudinal Study: A Biosocial Component, Oxfordshire A REC, Reference: 10/H0604/2).

#### English Longitudinal Study of Ageing (ELSA)

The English Longitudinal Study of Ageing is a longitudinal panel study of people aged 50 and over in England (10). The original sample of respondents was selected from households who had previously responded to the Health Surveys for England, a cross-sectional nationally representative household survey, in 1998, 1999 or 2001 (10). Data collection is conducted every two years by face-to-face interviews and paper self-completion surveys, with a nurse visit every second sweep (10). The sample is regularly replenished to ensure it remains representative of the over 50 population in England (10). More detail is available elsewhere (11).

##### Supplementary Table 5: ELSA data collection and sample achieved.^[4]^

| Sweep Number / Year | Interview type | Sample achieved | Nurse sample achieved | Sample replenished? |
| --- | --- | --- | --- | --- |
| 1  (2002-03) | Household questionnaire, individual interview, individual self-completion, cohort member assessment | 12,099 | - | - |
| 2  (2004-05) | Household questionnaire, individual interview, individual self-completion, cohort member assessment, nurse visit | 9,432 | 7,666 | - |
| 3  (2006-2007) | Household questionnaire, individual interview, individual self-completion, cohort member assessment | 9,771 | - | Y – aged 50-52 |
| 4  (2008-09) | Household questionnaire, individual interview, individual self-completion, cohort member assessment, nurse visit | 11,050 | 8,643 | Y – aged 50-74 |
| 5  (2010-11) | Household questionnaire, individual interview, individual self-completion, cohort member assessment | 10.274 | - | - |
| 6  (2012-13) | Household questionnaire, individual interview, individual self-completion, cohort member assessment, nurse visit | 10,601 | 8054 | Y- aged 50-55 |
| 7 (2014-15) | Household questionnaire, individual interview, individual self-completion, cohort member assessment | 9,666 | - | Y – aged 50-51 |
| 8 (2016-17) | Household questionnaire, individual interview, individual self-completion, cohort member assessment, nurse visit | 8,445 | 3525 | - |
| 9 (2018-19) | Household questionnaire, individual interview, individual self-completion, cohort member assessment | - | - | - |

ELSA was approved by the London Multicentre Research Ethics Committee (MREC/01/2/91), and informed consent was obtained from all participants.

### Sexual minority variable coding

In the main analyses respondents are recategorized as “Heterosexual” or “Sexual Minority”, however in subgroup analyses respondents are recategorized as “Heterosexual”, “Gay or Lesbian, “Bisexual”, “Other” and “Prefer Not To Say”.

Respondents were asked for the sexual identity label which best described them at age 17 in MCS (2018), wave 8 in ELSA (2016-17), at multiple waves in UKHLS, and at age 42 (2012) in BCS70. In UKHLS, BCS70 and ELSA, possible responses were as follows: “Heterosexual or Straight”, “Gay or Lesbian”, “Bisexual”, “Other”, Prefer Not to Say”, and “Don’t Know”.

In MCS, respondents who answered “Completely Heterosexual” was recoded as “Heterosexual”. Participants who answered “Completely Gay or Lesbian”, “Mainly Gay or Lesbian” and “Bisexual” are recoded as “Sexual Minority”. Respondents who answered “Mainly Heterosexual” or “Other” who also reported any same-sex attraction or no attraction are recoded as “Sexual Minority” with the remainder recoded as “Heterosexual”.

In Next Steps, respondents who answered “Heterosexual or Straight” are recoded as “Heterosexual”. Participants who answered “Gay or Lesbian”, “Bisexual”, or “Other” was recoded as “Sexual Minority”.

In UKHLS, respondents who answered “Heterosexual or Straight” are recoded as “Heterosexual”. Participants who answered “Gay or Lesbian”, “Bisexual”, or “Other” are recoded as “Sexual Minority”. As sexual identity is collected at multiple waves, the most recent response is used.

In BCS70, respondents who answered “Heterosexual or Straight” are recoded as “Heterosexual” and participants who answered “Gay or Lesbian”, or “Bisexual” are recoded as “Sexual Minority”.

In ELSA, respondents who answered “Heterosexual or Straight” are recoded as “Heterosexual” and participants who answered “Gay or Lesbian”, or “Bisexual” are recoded as “Sexual Minority”. Respondents who answered “Other” who also reported any same-sex attraction or no attraction are recoded as “Sexual Minority” with the remainder recoded as “Heterosexual”. Most recent response is used where multiple have been collected.

### Supplementary Figures and Tables

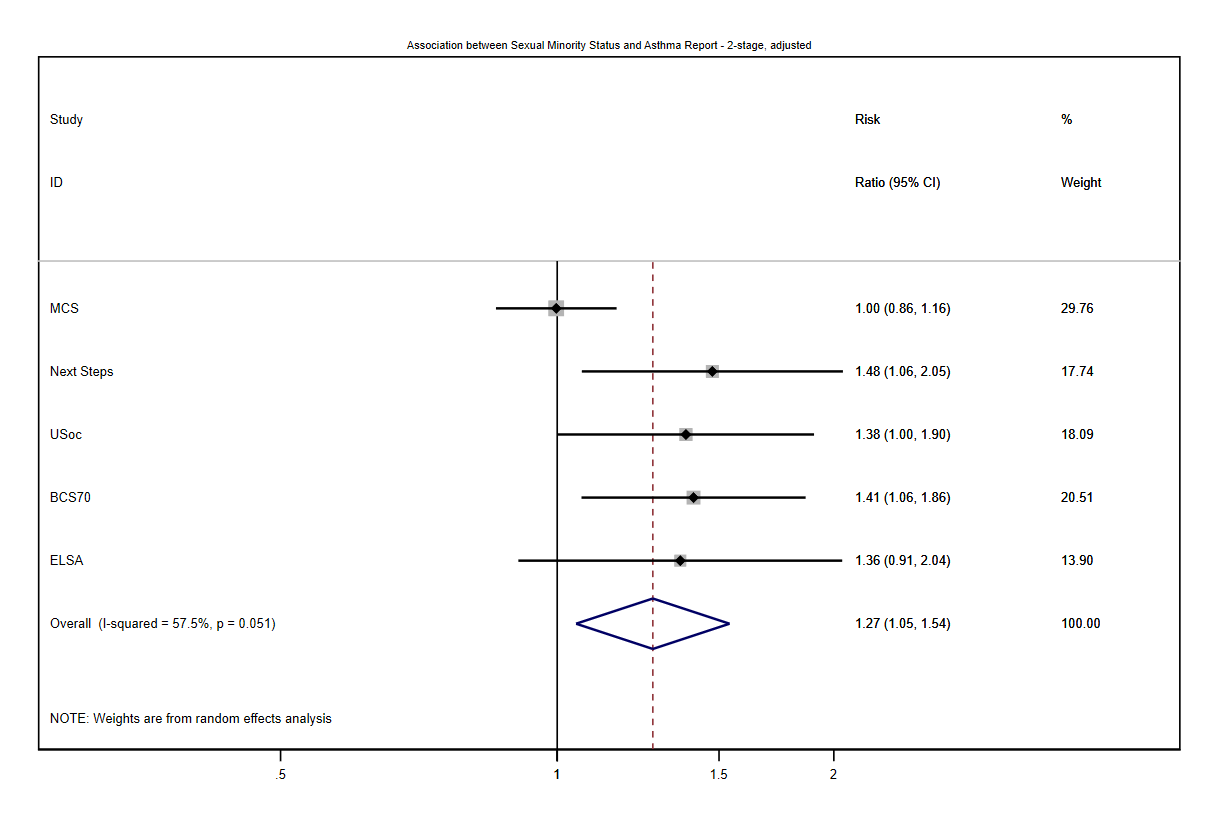

##### Supplementary Figure 1: Association between Sexual Minority status and Asthma report, meta-analysis adjusted risk ratios

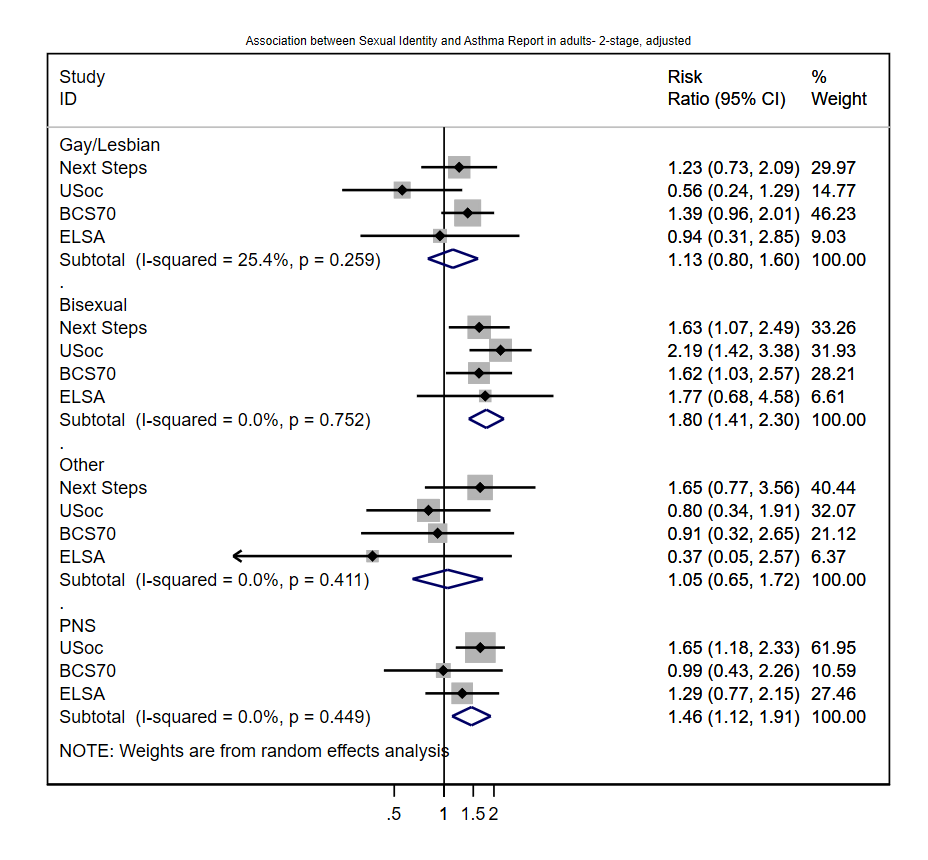

##### Supplementary Figure 2: Association between Sexual Identity status and Asthma report, meta-analysis adjusted risk ratios

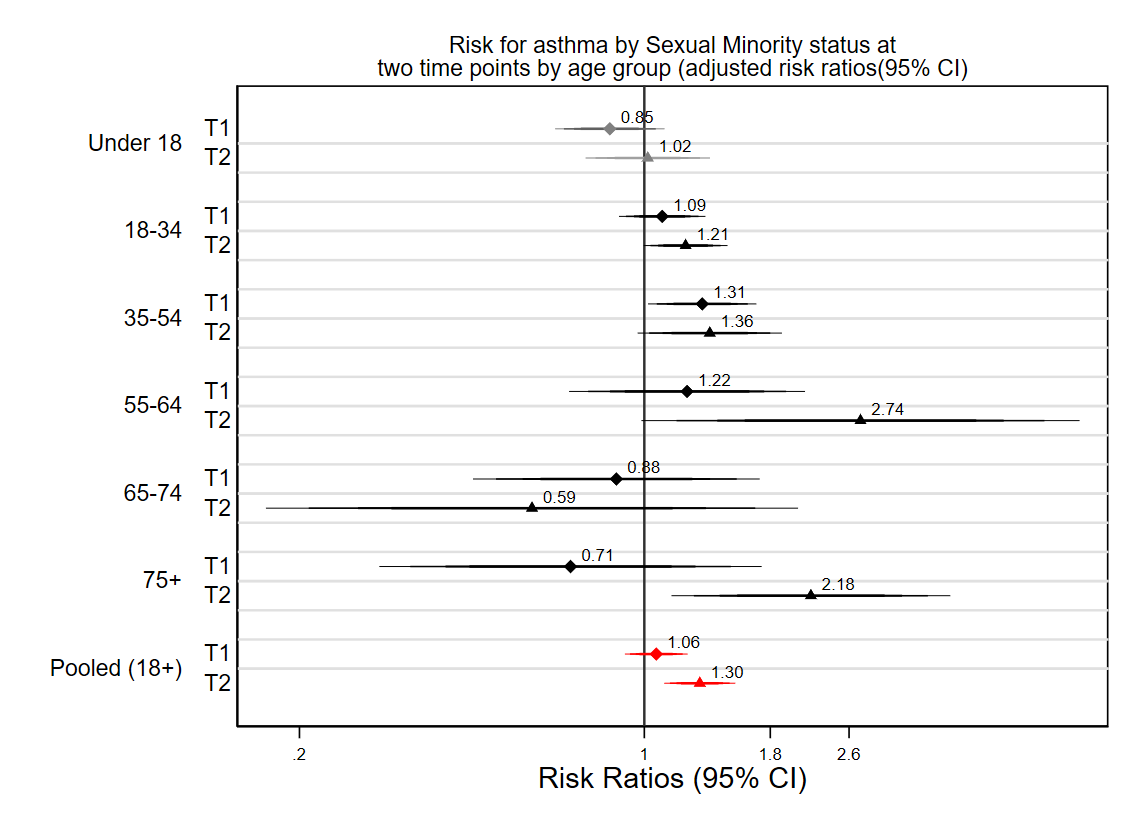

##### Supplementary Figure 3: Association between Sexual Minority status and Asthma report by study and age group across two timepoints, risk ratios

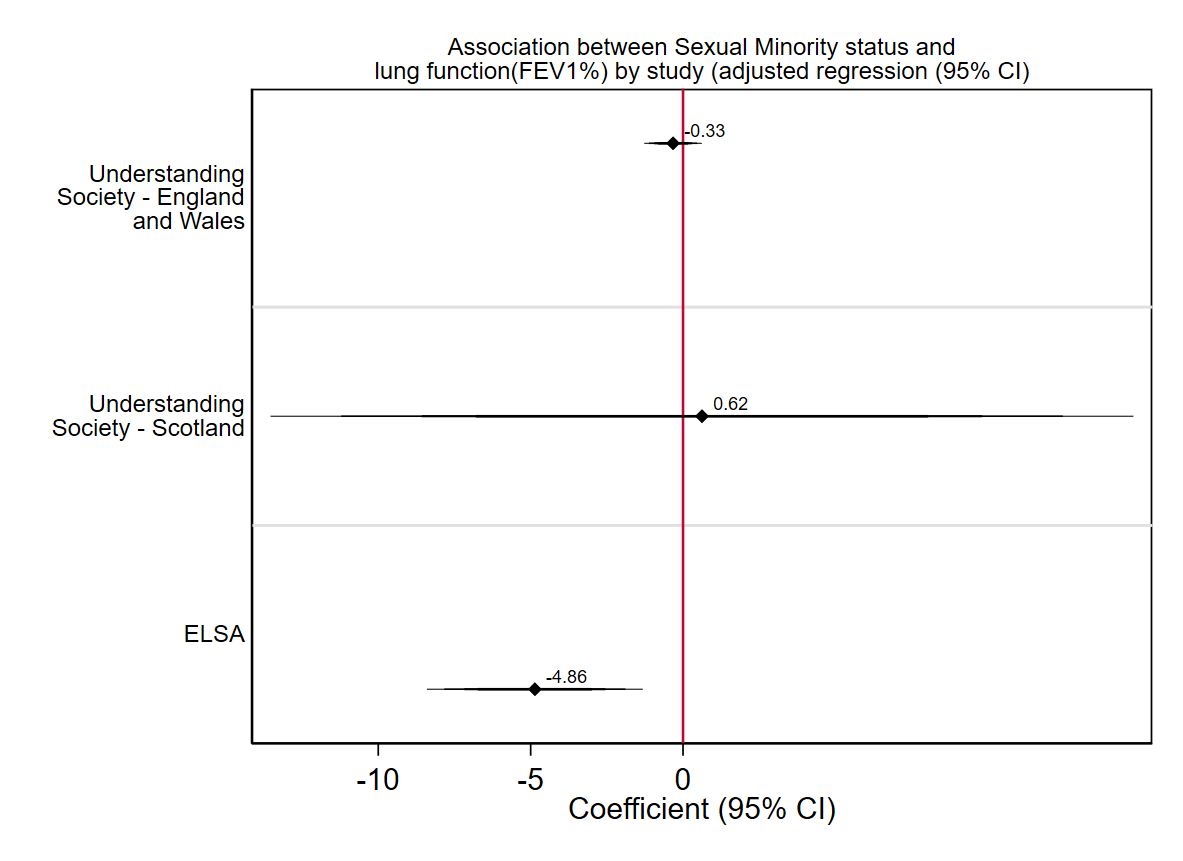

##### Supplementary Figure 4: Association between Sexual Minority status and lung function by continuous FEV1% by study, adjusted coefficients.

##### Supplementary Table 6: Asthma by sexual minority status in MCS, adjusted risk ratios (95% CI). Heterosexual as the reference category.

|  | Risk Ratio (95% CI) |
| --- | --- |
| Mainly Heterosexual categorised as Heterosexual | 1.02 (0.82 – 1.26) |
| Mainly Heterosexual categorised as Sexual Minority | 1.02 (0.87 -1.18) |

1. Sample description sourced from: (1,2)
2. Sample description sourced from:(5,6).
3. Sample description sourced from:(9).
4. Sample description sourced from: (10).
